# Can Generative Video Address the Clinical Video Data Gap? Evaluation of Synthetic Parkinsonian Hand Motions

**DOI:** 10.64898/2026.10.05.26363719

**Authors:** Yanbo Feng, Sriharsha Mopidevi, Kaleab Kinfu, Jean Park, Rene Vidal, Kevin B. Johnson

## Abstract

Computer vision approaches to disease motor assessment are limited by clinical video dataset scarcity and distributional imbalance, motivating interest in generative video models as a potential source of training data. We introduce a three-component framework for evaluating the clinical fidelity of synthetic medical motion data, assessing visual fidelity, biomechanical stability, and pathophysiological accuracy, and apply it to videos generated by the text-to-video model Sora 2 depicting Parkinsonian hand-fisting motions across MDS-UPDRS severity levels 0–2 (normal to mild–moderate motor impairment). We further use real-versus-real bootstrapping to compare synthetic-real differences against the natural variability of real clinical videos. Although generated videos appear visually realistic, quantitative analysis shows substantial divergence from real clinical recordings, including weaker severity-dependent patterns of bradykinesia and hypokinesia, less realistic tremor rhythms, and substantial temporal instability. These findings reveal a motion-domain gap between Sora-2-generated and real clinical motor behavior in this Parkinsonian hand-motion setting and demonstrate that visual realism alone is insufficient to establish the clinical utility of synthetic video data for training or evaluating medical computer vision systems.

## Introduction

The rapid advancement of text-to-video (T2V) generative models has sparked interest in using synthetic data for data augmentation to overcome the severe clinical data scarcity constraining medical AI due to strict privacy regulations. However, synthesizing specialized medical video represents a fundamental challenge in out-of-distribution (OOD) generation. While these general models excel at producing plausible everyday content, it remains unclear whether they can synthesize and augment OOD medical data with rigorous clinical validity. This raises a central question: can general-purpose generative models accurately synthesize clinically meaningful OOD medical motion video data?

To rigorously evaluate this OOD generative capacity, we utilize Parkinson’s disease (PD) motor assessment as a highly constrained, representative benchmark. PD is a neurodegenerative disorder affecting approximately 200 per 100,000 individuals [1]. While computer-vision frameworks have been proposed to objectively quantify PD kinematics [2, 3], their generalization is directly bottlenecked by the aforementioned lack of diverse training data. Consequently, standard clinical assessment still relies heavily on the subjective visual evaluation of symptoms like bradykinesia, rigidity, tremor, and hypokinesia [1, 4, 5] using established scales (e.g., UPDRS [6], and revised MDS-UPDRS [7]). This subjectivity contributes to documented misclassification rates of 15%–24% [8, 9, 10, 11], directly impacting critical interventions such as levodopa dosing and deep brain stimulation [12, 13]. Because authentic Parkinsonian motion requires strict adherence to complex pathophysiological rules, it provides an optimal stress test for evaluating whether general-purpose models can generate clinically valid synthetic video.

To evaluate general-purpose models’ clinical data synthesis ability, we introduce a physics-aware evaluation framework designed specifically for OOD medical motion synthesis. The framework integrates three complementary dimensions: visual fidelity, biomechanical stability, and pathophysiological accuracy, to systematically assess whether generated sequences merely appear realistic or actually follow the physiological rules underlying human movement. Applying this framework to Parkinsonian hand-fisting videos generated by Sora 2 reveals a substantial gap between visual plausibility and physiological validity, demonstrating why traditional generative evaluation metrics alone are insufficient for clinical applications.

## Related Works

### Hand Kinematics as a Rigorous Generative Benchmark

In the computer vision domain, human motion analysis increasingly relies on 3D joint keypoints and parametric models (e.g., SMPL [14]) to extract robust kinematics while preserving privacy. These representations have been successfully applied to PD macro-movements, such as gait classification [2] and gait severity grading [3]. However, fine-grained hand kinematics remain computationally under-explored due to their dense degrees of freedom, severe self-occlusions, and high biomechanical complexity [15]. This exact geometric complexity makes hand articulation, specifically when constrained by strict pathophysiological rules like tremor and finger bradykinesia [6, 7], an exceptionally rigorous stress test for OOD video synthesis. Concurrently, recent breakthroughs in robust 3D hand mesh and skeleton reconstruction [15, 16] provide a reliable mechanism for quantitative evaluation. In this study, we leverage these established reconstruction frameworks (i.e., MANO [15]) to map pixel-level generation into transparent, physics-based features, creating a grounded methodology to audit whether T2V models truly internalize complex OOD physical rules.

### Generative AI for Medical Data Augmentation

To address the scarcity of large-scale, diverse clinical datasets, studies have explored generative AI frameworks for data augmentation. This trend is visible across various medical modalities, including the synthesis of Electronic Health Records (EHR) [17] and static medical imaging [18]. In the realm of temporal human motion, recent work such as GAITGen [19] has investigated partially OOD synthetic data generation for Parkinsonian gait using healthy gait as input. The potential of these generative frameworks motivates our study. Specifically, the emergence of general-purpose T2V models raises an open question: to what extent can these broad-domain models perform visual clinical data synthesis? Because few papers have investigated the capacity of general-purpose T2V models to synthesize fine-grained neurological symptoms strictly from text prompts, our work serves as one of the initial exploratory steps to understand their current capabilities and limitations for medical video augmentation.

### Evaluation Paradigms for Generative Models

Assessing the quality of generative models introduces unique methodological challenges. Because evaluating OOD T2V generation lacks 1:1 paired ground-truth references, direct application of traditional pair-wise spatial error metrics is challenging. Instead, we adopt a commonly used standard composite evaluation strategy in image and video synthesis studies [20, 21] that leverages distributional metrics, including FID [22], FVD [23, 24], and FVMD [25], to observe broad visual and spatio-temporal fidelity. We complement these with reference-free, physics-based kinematic markers, including Power Spectral Density (PSD) for tremor analysis and distributional velocity metrics, to provide a preliminary profile of the pathophysiological characteristics of the synthetic motions.

## Methods

To systematically investigate the clinical meaningfulness of synthetic hand motion videos generated by general-purpose T2V models, we generated videos using Sora 2 and developed a population-comparing, physics-informed evaluation pipeline.

### Ethical Consideration

This study was deemed to be exempt by the Institutional Review Board of the University of Pennsylvania (Protocol #24-0286). The PD4T dataset was obtained under a formal data access agreement with Dr. Amirhossein Dadashzadeh and all data were de-identified prior to analysis.

### Real Data Acquisition

We utilized the restricted-access PD4T dataset [26]^1^, a clinical video archive designed for Parkinsonian action quality assessment. While PD4T contains 2,931 longitudinal RGB recordings, we isolated the 25 Frames-Per-Second (FPS) hand movement subset—comprising 848 videos (423 right-hand, 425 left-hand)—to evaluate fine-grained upper extremity kinematics during ten rapid, maximal-amplitude opening-and-closing cycles. Trained clinicians annotated this subset using the discrete MDS-UPDRS scale (0–4), spanning 234 UPDRS 0 (normal), 407 UPDRS 1 (slight), 179 UPDRS 2 (mild), 23 UPDRS 3 (moderate), and 5 UPDRS 4 (severe) recordings. These clinical scores serve as the ground-truth conditioning labels for our generative prompts and the baseline distributions for our evaluation pipeline.

### Real Data Pre-processing

To establish a robust baseline, 300 videos were randomly selected from the PD4T hand movement subset (UPDRS 0–2) with fixed random seed, stratified by severity and hand side (50 left/50 right per level). Levels 3 and 4 were excluded to maintain class balance and focus on early-stage symptom validation. For geometric consistency with Sora 2, videos were standardized to a 9:16 aspect ratio. Hand-centered cropping was performed via YOLO11n-pose-hands^2^ for alignment and de-identification, with facial features obscured via Gaussian blur using YOLOV11l-face^3^. Finally, the initial 25 frames were discarded to remove camera artifacts, and the first processed frame was extracted as the visual reference for generation.

### Prompt Generation Strategy

To generate synthetic videos with corresponding motor features in UPDRS levels 0, 1, and 2, we engineered one descriptive text prompt for each severity level, resulting in three prompts in total. To ensure strong correlation between the prompt and the corresponding severity, we performed (1) Clinical-to-Text Translation and (2) Prompt Architecture Design.

- *Clinical-to-Text Translation:* Since general-purpose T2V models lack inherent clinical comprehension, we utilized a Large Language Model (LLM) to assist in translating established MDS-UPDRS criteria and PD pathophysiology [27, 28] into explicit physical constraints. Clinical hallmarks like bradykinesia and hypokinesia were mapped to mechanical descriptors, such as moving through “thick glue” or fingers hitting an “invisible barrier,” to simulate progressive amplitude fatigue. Tremors were precisely instructed as “rhythmic, 5 Hz shaking” during motion pauses, effectively bridging clinical pathophysiology and generative T2V conditioning.
- *Prompt Architecture Design:* We engineered a standardized five-part template to scale motor symptoms systematically across all UPDRS severities (see Exemplar). [Visual Conditioning] anchored generative factors to the ground-truth reference image to mitigate hallucination. [Action] defined the global motor task, while [Timeline] dictated granular, beat-by-beat kinematic evolution. [Physics] applied environmental resistance to counteract the model’s bias toward temporal smoothing, and [Avoid] utilized negative constraints to suppress structural artifacts.

### Synthetic Video Generation

Conditioned on reference frames and the three T2V prompts, we generated the synthetic video set including 300 Sora-2-generated hand opening-and-closing videos. These videos have 10 seconds duration and 30 FPS with aspect ratio of 9:16. This frame rate is sufficient to capture the 4-6 Hz oscillatory patterns of PD tremor. This video set was balanced with 100 synthetic videos generated for each UPDRS class (0, 1, 2), creating 300 real-synthetic pairs for controlled kinematic comparison. Due to the balanced left and right-hand reference frames, the synthetic videos are also balanced for left and right hand classes.

### Data Processing and Interpolation

We resampled synthetic videos from 30 FPS to 25 FPS native rate of the real dataset to ensure temporal consistency. To account for the fixed 10-second duration of synthetic videos versus the variable durations of real video recordings, we randomly head-trimmed the synthetic videos to 8 to 10 seconds. We utilized the Hand Mesh Recovery (HaMeR) 3D reconstruction framework [16] to extract 3D joint keypoint sequences (*T*, 21, 3) and MANO [15] joint rotation (*θ*) sequences (*T*, 16, 3, 3), where T represents the temporal dimension of each video. To ensure data integrity, sequences with fewer than 70% valid reconstruction frames were discarded. Residual tracking gaps were mitigated using Akima interpolation [29] for the joint keypoints and Spherical Linear Interpolation (SLERP) [30] for the *θ* parameters.

### Evaluation Framework and Metrics

To provide a multidimensional assessment of videos generated by Sora 2, we propose our evaluation framework that quantitatively assesses (1) visual fidelity, (2) biomechanical consistency, and (3) pathophysiological accuracy.

### Evaluation Cohorts and Reconstruction Filtering

Different evaluation components used different analysis cohorts. Visual fidelity metrics including FID, FVD, and FVMD were computed on the full video cohorts, whereas HaMeR-dependent biomechanical and pathophysiological analyses were performed on the reconstruction-valid subset shown in Table 1. To assess potential reconstruction-domain differences between real and Sora 2-generated videos, we also summarized HaMeR reconstruction quality by source and UPDRS level, including 70% filter retention rate, valid-frame ratio, internal missing-frame burden, and maximum reconstruction gap.

**Table 1:** Analysis cohorts and HaMeR reconstruction-quality audit. Full *n* was used for FID, FVD, and FVMD; Valid *n* was used for HaMeR-dependent analyses after the 70% valid-frame filter. VFR denotes valid-frame ratio.

| UPDRS | Source | Full $n$ | Valid $n$ | VFR Med. [IQR] | Internal Missing Frames Med. | Maximum Gap Length Med. |
| --- | --- | --- | --- | --- | --- | --- |
| 0 | Real | 100 | 24 (24%) | 49.7% [29.9, 69.4] | 65.0 | 7.0 |
| 0 | Synthetic | 100 | 39 (39%) | 57.4% [21.3, 84.4] | 75.5 | 11.5 |
| 1 | Real | 100 | 41 (41%) | 60.4% [41.5, 88.6] | 68.5 | 7.0 |
| 1 | Synthetic | 100 | 44 (44%) | 61.4% [27.3, 85.1] | 81.0 | 13.0 |
| 2 | Real | 100 | 47 (47%) | 68.0% [48.0, 91.9] | 60.5 | 8.0 |
| 2 | Synthetic | 100 | 46 (46%) | 64.1% [33.6, 92.1] | 68.5 | 13.0 |

### Visual Fidelity Assessment

In this step, to quantify the domain gap between the real video population and the synthetic population, we employed Fréchet distance metrics, which assess the divergence between the multivariate Gaussian distributions (*N* (*µ*_*gt*_, Σ_*gt*_), *N* (*µ*_*gen*_, Σ_*gen*_)) of the feature embeddings of the ground-truth real videos and the generated videos:

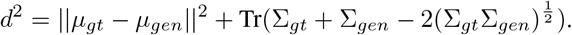

- *Fréchet Inception Distance (FID) [22]:* It evaluates the spatial realism and static visual quality of individual frames by computing Fréchet distance of image embeddings from a pre-trained Inception-v3 network.
- *Fréchet Video Distance (FVD) [23, 24]:* This metric evaluates broad spatio-temporal coherence and temporal fluency of the generated sequences by computing Fréchet distance between video embeddings from a pre-trained I3D network.
- *Fréchet Video Motion Distance (FVMD) [25]:* FVMD specifically evaluates physical plausibility by computing the Fréchet distance of fine-grained point trajectories extracted via a pre-trained PIPS++ point-tracking model.

### Biomechanical Assessment

To verify the physical and anatomical integrity of the generated hand structures, we extracted intrinsic biomechanical properties from the reconstructed 3D joint keypoints (*P*) and the MANO [15] parameter sequences (*θ*).

- *Rigid-Body Stability and Proportions:* Instead of simple variance, we computed the dimensionless Coefficient of Variation (*CV* = (*σ*_*L*_*/µ*_*L*_) *×* 100) for the Euclidean lengths (*L*) of all 20 hand bones across time. A high CV indicates “morphing” artifacts, representing a failure to maintain rigid-body constraints. Additionally, we calculated the anatomical length ratio between the index and pinky fingers to ensure the generated meshes preserve physiological human proportions.
- *Internal Joint Kinematics:* Leveraging the MANO [15] pose parameter (*θ*) sequences, we computed the relative rotational magnitudes (via axis-angle representations) for the internal hand joints. This isolates pure finger articulation to provide a direct comparison of the synthetic Range of Motion (ROM) against the physiological limits observed in the real-world baseline.

### Pathophysiological Assessment

To quantitatively audit the clinical realism of the synthesized PD motor symptoms, we extracted a suite of reference-free kinematic metrics from the keypoint trajectories and their derived velocity fields 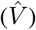.

- *Tremor and Frequency Domain (PSD):* We computed the Power Spectral Density (PSD) of the 3D middle fingertip trajectory using Welch’s method. To specifically quantify parkinsonian resting and action tremors, we integrated the spectral power within the 4–9 Hz band. The dominant peak frequency within this band was also extracted to verify rhythmic consistency against the real data distributions.
- *Bradykinesia and Sequence Effect:* To robustly evaluate movement slowness and progressive fatigue, we aggregated the velocity magnitude across all five fingertips. We extracted individual open-close movement cycles using a dynamic prominence heuristic. By fitting a linear trendline across the peak velocities (*v*_*peak*_) of consecutive cycles, we derived the fatigue slope, where a negative value explicitly quantifies the progressive sequence effect characteristic of PD.
- *Hypokinesia and Spatial Underscaling:* Hypokinesia is assessed by measuring the spatial scaling of the synthesized motions. Using the aggregated distance from the fingertips to the wrist, we defined the cycle amplitude (*A*_*cycle*_) as the difference between maximum extension and flexion. We computed both the absolute mean amplitude and an amplitude completeness ratio, defined as the ratio of the overall average amplitude to the initial baseline amplitude (derived from the first distinct cycles). These metrics evaluate the model’s capacity to simulate the spatial underscaling of muscle force.

Because fatigue slope and PSD metrics may be sensitive to outliers and reconstruction artifacts, we performed two additional interpretability checks. For fatigue slope, we inspected median and interquartile range summaries to determine whether large mean *±* SD values were driven by extreme trajectories. For PSD analysis, we inspected high-frequency spectral ratios to assess whether tremor-band differences reflected excess synthetic noise rather than tremor-band motion structure.

### Exploratory Downstream Severity Classification

To assess whether Sora 2-generated kinematic features provide useful downstream signal for clinical prediction, we trained feature-based UPDRS severity classifiers using the extracted reconstruction-dependent biomechanical and pathophysiological metrics. We compared three training regimes: real-only, synthetic-only, and real-plus-synthetic training. All models were evaluated on held-out real clinical videos using repeated stratified splits. Because the goal was to evaluate downstream transfer of extracted motion features rather than to develop a new classifier, we used class-balanced multinomial logistic regression with median imputation and feature standardization. Performance was summarized using balanced accuracy, macro-F1, and ordinal mean absolute error.

## Experiments

### Settings

All experiments used 300 real and 300 Sora 2-generated videos balanced across UPDRS 0–2. Visual-fidelity metrics were computed on the full cohorts, whereas reconstruction-dependent metrics used the HaMeR-valid subset described in Table 1.

### Evaluation

Visual fidelity (FID [22], FVD [23, 24], FVMD [25]) was assessed using an iterative bootstrapping protocol (*T* = 1000) to ensure statistical rigor. In each iteration, ground-truth clinical embeddings were partitioned into non-overlapping subsets to establish a Real-vs-Real (RvR) baseline of intrinsic variance, which was then compared against the Real-vs-Fake (RvF) distance. This RvR baseline provided an empirical estimate of natural clinical-video variability, allowing RvF discrepancies to be interpreted relative to real-data variation rather than as uncalibrated absolute distances. This framework yielded robust mean scores and 95% Confidence Intervals (CI). Biomechanical stability was quantified by the Coefficient of Variation (CV) across 20 rigid bone lengths, anatomical Index-to-Pinky proportions, and joint Range of Motion (ROM). Pathophysiological realism was profiled through signal processing of kinematic trajectories, specifically: 3D velocity magnitudes to derive fatigue slopes, dynamic prominence extraction for amplitude completeness, and 4–9 Hz Power Spectral Density (PSD) integration to isolate tremor power. Results are reported as Mean *±* SD to compare synthetic generations against clinical baselines.

## Results

In this section, we present our quantitative findings across three distinct evaluation pillars: visual fidelity, biomechanical stability, and pathophysiological accuracy.

### Visual Fidelity

Visual fidelity was assessed using iterative bootstrapping to compare intrinsic Real-vs-Real (RvR) variability with the Real-vs-Fake (RvF) generation gap. As detailed in Table 2, FID showed a consistent appearance gap between real and Sora 2-generated videos across all UPDRS levels, despite tight RvR baselines. FVD indicated that short-context (*FV D*_16_) temporal coherence was closer to real-video variability, whereas longer-context (*FV D*_128_) evaluation amplified the synthetic-real divergence; stride-8 downsampling reduced *FV D*_128_ but did not eliminate this synthetic-real divergence. The largest discrepancy appeared in FVMD, where RvF motion-trajectory distances were orders of magnitude larger than RvR baselines across all severities. These findings indicate that Sora 2-generated videos can appear visually plausible while exhibiting substantial motion-domain divergence.

**Table 2:** Visual Fidelity Metrics across UPDRS Severity Levels. The evaluation utilizes an Iterative Bootstrapping approach (*T* = 1000) to establish both the intrinsic Real-vs-Real (RvR) intra-distributional baseline and the Real-vs-Fake (RvF) generation gap.

| Severity | Metric | Iterative Bootstrapping ($T = 1000$) | | | |
| --- | --- | --- | --- | --- | --- |
|  |  | RvR Mean | RvR 95% CI | RvF Mean | RvF 95% CI |
| UPDRS 0 | FID | 2.03 | [1.97, 2.11] | 21.45 | [21.10, 21.78] |
| | $FVD_{16}$ | 237.52 | [216.70, 271.11] | 335.88 | [300.59, 373.90] |
| | $FVD_{128}$ | 350.86 | [302.20, 431.68] | 783.97 | [662.87, 915.25] |
| | $FVD_{128}$ (Stride 8) | 208.70 | [189.06, 236.77] | 306.54 | [272.25, 345.15] |
|  | FVMD | 177.77 | [167.53, 192.06] | 7865.44 | [7693.76, 8028.96] |
| UPDRS 1 | FID | 1.73 | [1.67, 1.80] | 21.61 | [21.33, 21.91] |
| | $FVD_{16}$ | 254.91 | [234.69, 284.12] | 324.84 | [289.67, 363.84] |
| | $FVD_{128}$ | 363.15 | [323.39, 414.93] | 567.08 | [496.86, 649.05] |
| | $FVD_{128}$ (Stride 8) | 220.29 | [201.57, 247.05] | 279.18 | [253.82, 306.15] |
|  | FVMD | 149.48 | [143.51, 156.76] | 5678.10 | [5550.82, 5794.94] |
| UPDRS 2 | FID | 1.29 | [1.25, 1.35] | 21.98 | [21.74, 22.22] |
| | $FVD_{16}$ | 237.90 | [217.03, 268.11] | 324.73 | [285.13, 365.74] |
| | $FVD_{128}$ | 355.45 | [310.63, 422.88] | 558.37 | [491.62, 638.81] |
| | $FVD_{128}$ (Stride 8) | 220.39 | [201.61, 248.11] | 298.59 | [263.98, 339.15] |
|  | FVMD | 93.40 | [88.99, 98.16] | 8389.18 | [8219.71, 8545.82] |

### Biomechanical Stability

Unlike the Fréchet-distance visual-fidelity metrics, biomechanical and pathophysiological analyses required valid HaMeR-derived 3D keypoints and MANO pose sequences and were therefore performed on the reconstruction-valid subset described in Table 1. Synthetic videos were not systematically retained at lower rates than real videos after the 70% valid-frame filter, although they showed larger internal missing-frame burden and longer maximum reconstruction gaps, suggesting greater temporal fragmentation in reconstructed trajectories. As detailed in Table 3, Sora 2-generated videos showed broadly comparable bone-length CV, anatomical proportions, absolute hand size, and mean joint ROM to real baselines across UPDRS levels. Overall, Sora 2 preserved macroscopic hand shape and joint articulation more successfully than subtle disease-dependent biomechanical trends.

**Table 3:**
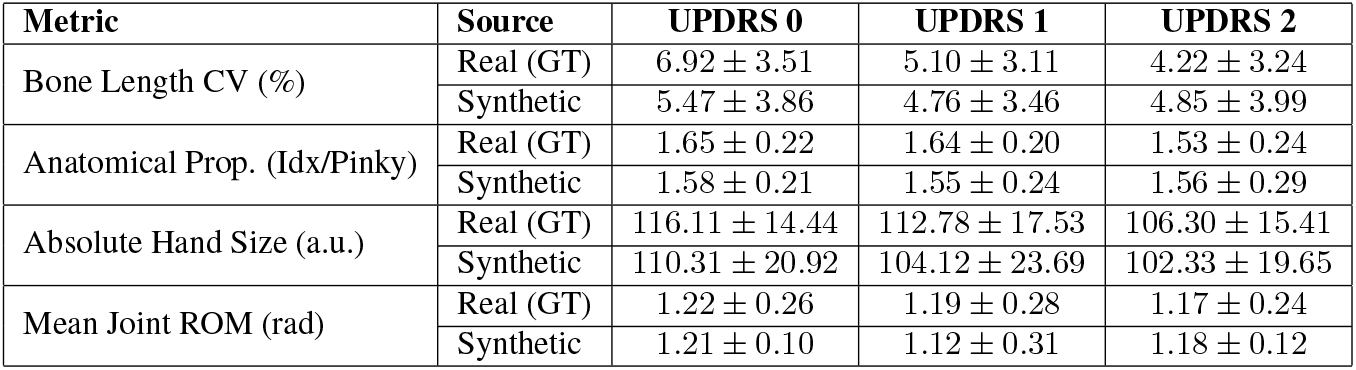
Quantitative Biomechanical Assessment. The table compares explicit 3D rigid-body kinematics and anatomical proportions extracted from real clinical baselines (GT) against synthetic Sora 2 generations across UPDRS severities. Values are reported as Mean *±* Standard Deviation.

| Metric | Source | UPDRS 0 | UPDRS 1 | UPDRS 2 |
| --- | --- | --- | --- | --- |
| Bone Length CV (%) | Real (GT) | 6.92 ± 3.51 | 5.10 ± 3.11 | 4.22 ± 3.24 |
| | Synthetic | 5.47 ± 3.86 | 4.76 ± 3.46 | 4.85 ± 3.99 |
| Anatomical Prop. (Idx/Pinky) | Real (GT) | 1.65 ± 0.22 | 1.64 ± 0.20 | 1.53 ± 0.24 |
| | Synthetic | 1.58 ± 0.21 | 1.55 ± 0.24 | 1.56 ± 0.29 |
| Absolute Hand Size (a.u.) | Real (GT) | 116.11 ± 14.44 | 112.78 ± 17.53 | 106.30 ± 15.41 |
| | Synthetic | 110.31 ± 20.92 | 104.12 ± 23.69 | 102.33 ± 19.65 |
| Mean Joint ROM (rad) | Real (GT) | 1.22 ± 0.26 | 1.19 ± 0.28 | 1.17 ± 0.24 |
| | Synthetic | 1.21 ± 0.10 | 1.12 ± 0.31 | 1.18 ± 0.12 |

### Pathophysiological Accuracy

Quantitative Parkinsonian motor markers are summarized in Table 4 and visualized in Figures 1–2. Real videos showed severity-dependent reductions in peak velocity and movement amplitude across UPDRS 0–2, whereas Sora 2-generated videos remained in comparable numerical ranges but failed to reproduce a consistent progressive severity gradient. Synthetic fatigue slopes showed much larger variance than real slopes; robust median/IQR summaries in Table 5 indicated that this variance was partly driven by extreme trajectories rather than a consistent shift in central tendency. Representative synthetic sequences in Figure 1 showed irregular cycle timing, missing or poorly detected cycles, and mid-sequence arrests, suggesting trajectory instability rather than physiologically structured fatigue progression. As displayed in Figure 2, frequency-domain analysis showed lower 4–9 Hz tremor-band power and lower-frequency clustering in synthetic videos. Because high-frequency ratios were not elevated in synthetic videos, shown in Table 6, these PSD differences were unlikely to be explained simply by excess high-frequency noise and instead suggest incomplete reproduction of real tremor-band motion structure. differences were driven by excess high-frequency synthetic noise, while tremor fraction summarizes the proportion of spectral power concentrated in the 4–9 Hz tremor band.

**Table 4:**
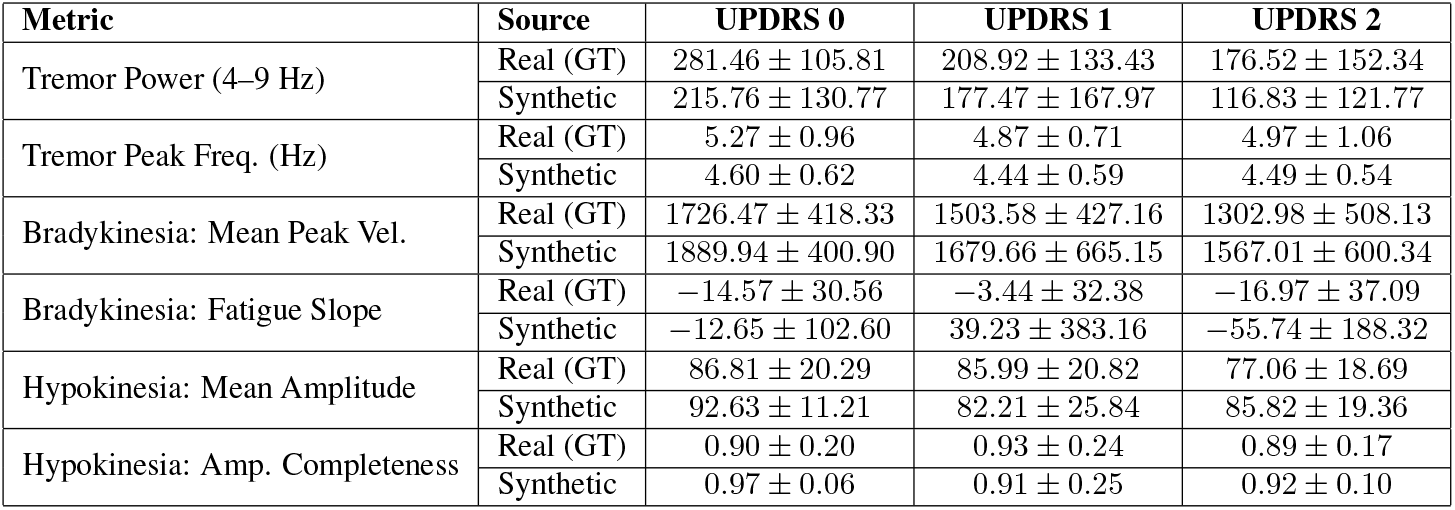
Quantitative Pathophysiological Assessment. The table evaluates the clinical realism of generated Parkinsonian motor symptoms by comparing kinematic markers of tremor, bradykinesia, and hypokinesia between real baselines and synthetic generations. Values are reported as Mean *±* Standard Deviation.

| Metric | Source | UPDRS 0 | UPDRS 1 | UPDRS 2 |
| --- | --- | --- | --- | --- |
| Tremor Power (4–9 Hz) | Real (GT) | 281.46 ± 105.81 | 208.92 ± 133.43 | 176.52 ± 152.34 |
| | Synthetic | 215.76 ± 130.77 | 177.47 ± 167.97 | 116.83 ± 121.77 |
| Tremor Peak Freq. (Hz) | Real (GT) | 5.27 ± 0.96 | 4.87 ± 0.71 | 4.97 ± 1.06 |
| | Synthetic | 4.60 ± 0.62 | 4.44 ± 0.59 | 4.49 ± 0.54 |
| Bradykinesia: Mean Peak Vel. | Real (GT) | 1726.47 ± 418.33 | 1503.58 ± 427.16 | 1302.98 ± 508.13 |
| | Synthetic | 1889.94 ± 400.90 | 1679.66 ± 665.15 | 1567.01 ± 600.34 |
| Bradykinesia: Fatigue Slope | Real (GT) | −14.57 ± 30.56 | −3.44 ± 32.38 | −16.97 ± 37.09 |
| | Synthetic | −12.65 ± 102.60 | 39.23 ± 383.16 | −55.74 ± 188.32 |
| Hypokinesia: Mean Amplitude | Real (GT) | 86.81 ± 20.29 | 85.99 ± 20.82 | 77.06 ± 18.69 |
| | Synthetic | 92.63 ± 11.21 | 82.21 ± 25.84 | 85.82 ± 19.36 |
| Hypokinesia: Amp. Completeness | Real (GT) | 0.90 ± 0.20 | 0.93 ± 0.24 | 0.89 ± 0.17 |
| | Synthetic | 0.97 ± 0.06 | 0.91 ± 0.25 | 0.92 ± 0.10 |

**Table 5:**
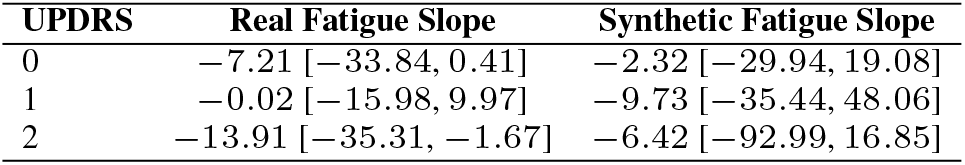
Robust summary of fatigue-slope distributions. Values are median [IQR]. This table complements the mean *±* SD summaries in Table 4 and shows whether large standard deviations are driven by broad central shifts or extreme trajectories.

| UPDRS | Real Fatigue Slope | Synthetic Fatigue Slope |
| --- | --- | --- |
| 0 | −7.21 [−33.84, 0.41] | −2.32 [−29.94, 19.08] |
| 1 | −0.02 [−15.98, 9.97] | −9.73 [−35.44, 48.06] |
| 2 | −13.91 [−35.31, −1.67] | −6.42 [−92.99, 16.85] |

**Table 6:** PSD artifact and tremor-band checks. Values are median [IQR]. High-frequency ratio was inspected to assess whether PSD differences were driven by excess high-frequency synthetic noise, while tremor fraction summarizes the proportion of spectral power concentrated in the 4−9 Hz tremor band.

| UPDRS | Source | High-Freq. Ratio | Tremor Fraction |
| --- | --- | --- | --- |
| 0 | Real | 0.0176 [0.0130, 0.0333] | 0.0917 [0.0697, 0.1133] |
| 0 | Synthetic | 0.0123 [0.0085, 0.0189] | 0.0626 [0.0476, 0.0848] |
| 1 | Real | 0.0125 [0.0055, 0.0233] | 0.0702 [0.0278, 0.1078] |
| 1 | Synthetic | 0.0119 [0.0062, 0.0233] | 0.0593 [0.0328, 0.0956] |
| 2 | Real | 0.0137 [0.0053, 0.0232] | 0.0574 [0.0230, 0.1018] |
| 2 | Synthetic | 0.0089 [0.0051, 0.0152] | 0.0429 [0.0318, 0.0671] |

**Figure 1:**
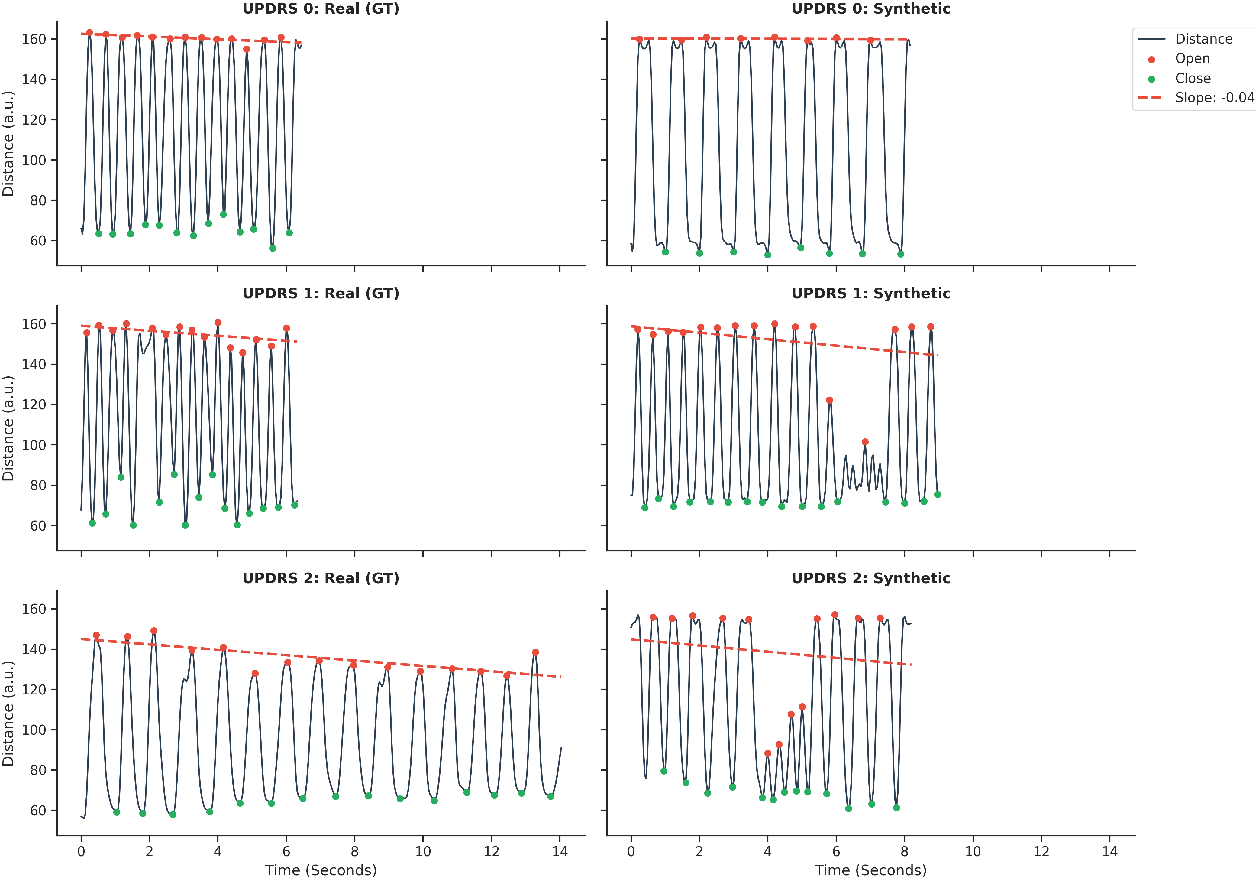
Temporal sequence analysis. Representative real sequences show bounded, regular open-close cycles, whereas Sora 2-generated examples show irregular timing, missing cycles, and mid-sequence arrests.

**Figure 2:**
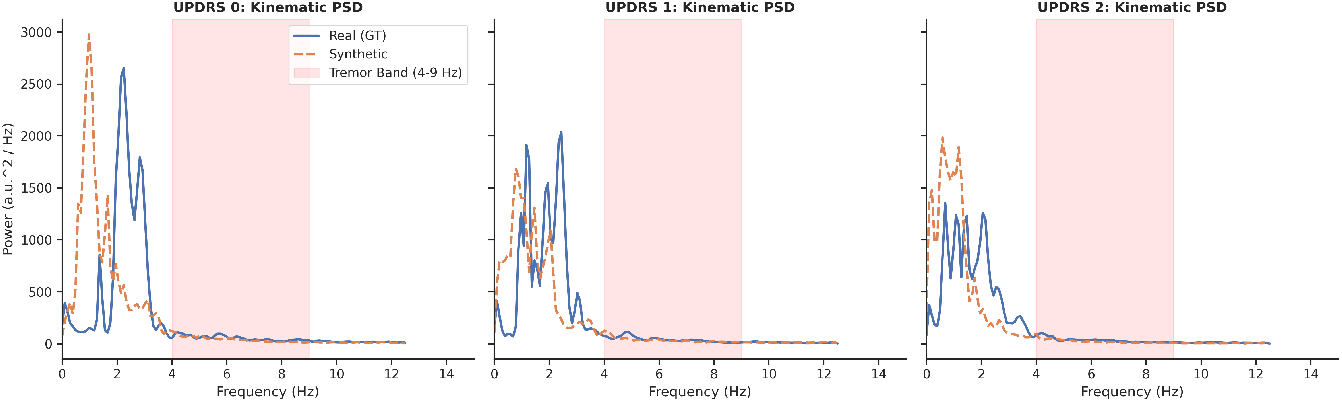
Kinematic power spectral density (PSD). Real sequences show stronger and more varied 4–9 Hz tremor-band structure, whereas Sora 2-generated sequences show lower spectral power and lower-frequency clustering.

### Exploratory Downstream Classification

As shown in Table 7, real-only training achieved the best held-out real-video UPDRS classification performance, whereas synthetic-only training showed poorer transfer to real videos. Adding Sora 2-generated samples to real training data did not improve performance and instead reduced balanced accuracy and macro-F1 relative to real-only training. These results suggest that Sora 2-generated kinematic features do not yet provide reliable augmentation signal for real-video UPDRS severity prediction.

**Table 7:**
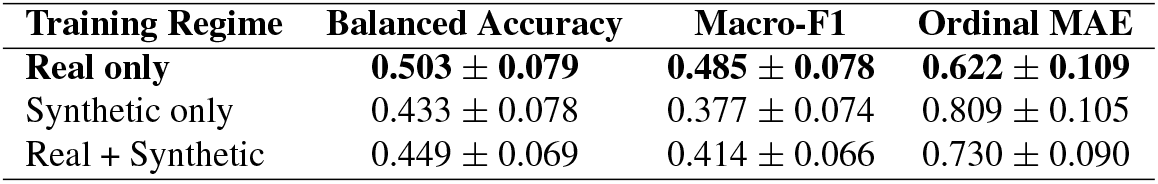
Exploratory downstream UPDRS severity classification on held-out real clinical videos using skeleton-derived biomechanical and pathophysiological features. Values are mean *±* SD across repeated stratified splits.

| Training Regime | Balanced Accuracy | Macro-F1 | Ordinal MAE |
| --- | --- | --- | --- |
| **Real only** | **0.503 ± 0.079** | **0.485 ± 0.078** | **0.622 ± 0.109** |
| Synthetic only | 0.433 ± 0.078 | 0.377 ± 0.074 | 0.809 ± 0.105 |
| Real + Synthetic | 0.449 ± 0.069 | 0.414 ± 0.066 | 0.730 ± 0.090 |

## Discussion

Our work advances clinical motion analysis by shifting the focus from developing generative models to evaluating their physiological realism. While prior studies model pathological dynamics using paired or task-specific motion data [19, 31], we introduce a reference-free, population-level framework for auditing the biomechanical and pathophysiological validity of generated clinical videos. A central strength of this framework is the real-versus-real bootstrapped baseline, which estimates natural variability among independent real clinical videos before judging synthetic-versus-real divergence. Applied to Sora 2, this framework exposes failure modes that are only partially captured by conventional visual metrics: generated videos may appear photorealistic and locally coherent, yet still exhibit substantial motion-domain divergence when evaluated through trajectory-based and clinical-motion features. Biomechanically, Sora 2-generated videos preserve macroscopic hand anatomy, including stable hand structure, plausible proportions, and reasonable joint articulation, but do not fully reproduce subtle disease-dependent mechanical patterns. Patho-physiologically, the limitations are more pronounced: although synthetic sequences can reach numerical ranges similar to real videos for some kinematic markers, they fail to reproduce progressive Parkinsonian severity gradients in bradykinesia, hypokinesia, fatigue, and tremor-band structure. These deviations suggest that Sora 2 approximates the superficial visual appearance of impairment rather than the underlying neuromotor mechanisms that govern real Parkinsonian motion. More broadly, clinical motor behaviors are rare out-of-distribution phenomena governed by physiological constraints that differ from general internet-video statistics. Therefore, synthetic clinical videos may appear visually realistic while violating properties required for downstream medical AI applications, underscoring the need for domain-aware evaluation frameworks before such data are used for model training, benchmarking, or augmentation.

### Limitations and Future Work

Several limitations should be considered. First, this study focused on Parkinsonian hand-fisting as a stringent stress test; future work should examine other clinical motion domains, such as gait analysis and stroke movement assessment in adults. Second, monocular 3D hand reconstruction introduces estimation error. Although applying the same HaMeR pipeline to real and Sora 2-generated videos improves population-level comparability, HaMeR may respond differently to synthetic textures, lighting, hand-boundary artifacts, and temporally fragmented motion. Our reconstruction-quality audit showed that Sora 2-generated videos were not systematically retained at lower rates than real videos after the 70% valid-frame filter, but they showed larger internal missing-frame burden and longer reconstruction gaps. The 70% filter may also introduce non-random exclusion if occlusion, motion blur, irregular finger motion, tremor, or severe bradykinesia reduce reconstruction validity; therefore, reconstruction-dependent findings should be interpreted within the HaMeR-valid subset. Finally, this study evaluated Sora 2 specifically; future work should compare additional general-purpose, open-source, and domain-adapted video generators and systematically assess prompt sensitivity.

## Conclusion

Our multidimensional framework offers a structured method to audit generative medical videos before machine-learning deployment by shifting evaluation from visual coherence alone to physiological validity. Although Sora 2-generated videos present anatomically plausible hand structures, they fail to reproduce key neuromotor patterns characteristic of Parkinsonian impairment, including clinically meaningful kinematic and pathophysiological dynamics. These findings show that evaluating synthetic clinical videos solely on visual realism risks embedding hidden clinical distortions into downstream training pipelines, underscoring the need for domain-aware evaluation frameworks to benchmark future clinical motion synthesis.

## Data Availability

The real clinical videos analyzed in this study were obtained from the PD4T dataset through its controlled-access data request process and cannot be redistributed by the authors. Access to PD4T may be requested from the original dataset maintainers in accordance with their data access procedures. Synthetic videos generated for this study and derived quantitative data are available from the authors upon reasonable request. Summary data supporting the findings of this study are included in the manuscript and its supplementary materials.

https://github.com/Plrbear/PECoP

## Acknowledgments

This work was funded by the NIH Pioneer Award, #DP1LM014558. The authors would like to thank the creators and maintainers of the PD4T dataset for providing the clinical video archive used in this study. We also acknowledge OpenAI’s Sora 2, Databricks computational infrastructure, and the open-source tools HaMeR [16], YOLO11n-pose-hands, and YOLOV11l-face for supporting video generation, 3D kinematic extraction, and preprocessing.

## Footnotes

1 Access to the PD4T dataset can be requested at: https://github.com/Plrbear/PECoP.git

2 *YOLO11n-pose-hands* is available at: https://github.com/chrismuntean/YOLO11n-pose-hands.git

3x *YOLOV11l-face* is available at: https://github.com/akanametov/yolo-face.git

## References

1. Parkinson I, Society MD. Parkinson’s Disease & Parkinsonism; 2025. Available from: https://www.movementdisorders.org/MDS/About/Movement-Disorder-Overviews/Parkinsons-Disease--Parkinsonism.htm.

2. Lu M, Poston K, Pfefferbaum A, Sullivan EV, Fei-Fei L, Pohl KM, et al. Vision-based estimation of MDS-UPDRS gait scores for assessing Parkinson’s disease motor severity. In: International Conference on Medical Image Computing and Computer-Assisted Intervention. Springer; 2020. p. 637–47.

3. Ben Hadj-Alouane N, Dhoot A, Turki-Hadj Alouane M, Pangracious V. Severity Classification of Parkinson’s Disease via Synthesis of Energy Skeleton Images from Videos Produced in Uncontrolled Environments. Diagnostics. 2024;14(23):2685.

4. Bloem BR, Okun MS, Klein C. Parkinson’s disease. The Lancet. 2021;397(10291):2284–303.

5. Kassubek J. Diagnostic procedures during the course of Parkinson’s disease. Basal Ganglia. 2014;4(1):15–8.

6. Fahn S. Unified Parkinson’s disease rating scale. Recent developments in Parkinson’s disease. 1987:153–63.

7. Goetz CG, Tilley BC, Shaftman SR, Stebbins GT, Fahn S, Martinez-Martin P, et al. Movement Disorder Society-sponsored revision of the Unified Parkinson’s Disease Rating Scale (MDS-UPDRS): scale presentation and clinimetric testing results. Movement disorders: official journal of the Movement Disorder Society. 2008;23(15):2129–70.

8. Tolosa E, Garrido A, Scholz SW, Poewe W. Challenges in the diagnosis of Parkinson’s disease. The Lancet Neurology. 2021;20(5):385–97.

9. Rajput AH, Rajput A. Accuracy of Parkinson disease diagnosis unchanged in 2 decades. Lippincott Williams & Wilkins Hagerstown, MD; 2014.

10. Schrag A, Ben-Shlomo Y, Quinn N. How valid is the clinical diagnosis of Parkinson’s disease in the community? Journal of Neurology, Neurosurgery & Psychiatry. 2002;73(5):529–34.

11. Hughes AJ, Daniel SE, Kilford L, Lees AJ. Accuracy of clinical diagnosis of idiopathic Parkinson’s disease: a clinico-pathological study of 100 cases. Journal of neurology, neurosurgery & psychiatry. 1992;55(3):181–4.

12. Paoletti FP, Tambasco N, Parnetti L. Levodopa treatment in Parkinson’s disease: earlier or later? Annals of translational medicine. 2019;7(Suppl 6):S189.

13. Artusi CA, Lopiano L, Morgante F. Deep brain stimulation selection criteria for Parkinson’s disease: time to go beyond CAPSIT-PD. Journal of clinical medicine. 2020;9(12):3931.

14. Loper M, Mahmood N, Romero J, Pons-Moll G, Black MJ. SMPL: A skinned multi-person linear model. In: Seminal Graphics Papers: Pushing the Boundaries, Volume 2; 2023. p. 851–66.

15. Romero J, Tzionas D, Black MJ. Embodied hands: Modeling and capturing hands and bodies together. arXiv preprint 220102610. 2022.

16. Pavlakos G, Shan D, Radosavovic I, Kanazawa A, Fouhey D, Malik J. Reconstructing Hands in 3D with Transformers. In: CVPR; 2024.

17. Choi E, Biswal S, Malin B, Duke J, Stewart WF, Sun J. Generating multi-label discrete patient records using generative adversarial networks. In: Machine learning for healthcare conference. PMLR; 2017. p. 286–305.

18. Frid-Adar M, Diamant I, Klang E, Amitai M, Goldberger J, Greenspan H. GAN-based synthetic medical image augmentation for increased CNN performance in liver lesion classification. Neurocomputing. 2018;321:321–31.

19. Adeli V, Mehraban S, Mirmehdi M, Whone A, Filtjens B, Dadashzadeh A, et al. GAITGen: Disentangled Motion-Pathology Impaired Gait Generative Model–Bringing Motion Generation to the Clinical Domain. In: Proceedings of the IEEE/CVF Winter Conference on Applications of Computer Vision; 2026. p. 3150–61.

20. Huang Z, He Y, Yu J, Zhang F, Si C, Jiang Y, et al. Vbench: Comprehensive benchmark suite for video generative models. In: Proceedings of the IEEE/CVF Conference on Computer Vision and Pattern Recognition; 2024. p. 21807–18.

21. Ho J, Chan W, Saharia C, Whang J, Gao R, Gritsenko A, et al. Imagen video: High definition video generation with diffusion models. arXiv preprint 221002303. 2022.

22. Heusel M, Ramsauer H, Unterthiner T, Nessler B, Hochreiter S. Gans trained by a two time-scale update rule converge to a local nash equilibrium. Advances in neural information processing systems. 2017;30.

23. Unterthiner T, Van Steenkiste S, Kurach K, Marinier R, Michalski M, Gelly S. Towards accurate generative models of video: A new metric & challenges. arXiv preprint 181201717. 2018.

24. Unterthiner T, Van Steenkiste S, Kurach K, Marinier R, Michalski M, Gelly S. FVD: A new metric for video generation. 2019.

25. Liu J, Qu Y, Yan Q, Zeng X, Wang L, Liao R. Fréchet Video Motion Distance: A Metric for Evaluating Motion Consistency in Videos. arXiv preprint 240716124. 2024.

26. Dadashzadeh A, Duan S, Whone A, Mirmehdi M. Pecop: Parameter efficient continual pretraining for action quality assessment. In: Proceedings of the IEEE/CVF Winter Conference on applications of computer vision; 2024. p. 42–52.

27. Berardelli A, Rothwell JC, Thompson PD, Hallett M. Pathophysiology of bradykinesia in Parkinson’s disease. Brain. 2001;124(11):2131–46.

28. Dirkx MF, Bologna M. The pathophysiology of Parkinson’s disease tremor. Journal of the Neurological Sciences. 2022;435:120196.

29. Akima H. A new method of interpolation and smooth curve fitting based on local procedures. Journal of the ACM (JACM). 1970;17(4):589–602.

30. Shoemake K. Animating rotation with quaternion curves. In: Proceedings of the 12th annual conference on Computer graphics and interactive techniques; 1985. p. 245–54.

31. Kocabas M, Athanasiou N, Black MJ. Vibe: Video inference for human body pose and shape estimation. In: Proceedings of the IEEE/CVF conference on computer vision and pattern recognition; 2020. p. 5253–63.

